# Genetic influence of BCAA metabolism on type 2 diabetes and coronary artery disease, independent of traditional risk factors

**DOI:** 10.64898/2026.03.28.26349580

**Authors:** Shota Nakamura, Masaru Koido, Yunye He, Yohei Takeuchi, Hiromi Tsuru, Yoji Sagiya, Akiko Nagai, Takayuki Morisaki, Koichi Matsuda, Yoichiro Kamatani

## Abstract

Type 2 diabetes (T2D) and cardiovascular disease are major global health burdens. Branched-chain amino acid (BCAA) metabolism has been implicated as a potential therapeutic target, but it remains unclear whether its associations with disease risk are independent of traditional risk factors such as obesity and dyslipidemia. We leveraged genomic structural equation modeling of large-scale genome-wide association studies (GWAS) from European and East Asian populations, including the largest East Asian GWAS of BCAA levels (n = 42,826). We identified a genetic factor influencing BCAA metabolism independently of body mass index and circulating lipid levels. A cross-population polygenic score derived from this factor was associated with hemoglobin A1c, blood glucose, and the onset of both T2D and coronary artery disease. These findings provide the first genetic insight in humans that BCAA metabolism is involved in T2D and CAD beyond traditional risk factors, highlighting its clinical relevance and therapeutic potential.

## Introduction

Type 2 diabetes (T2D) and cardiovascular disease (CVD) pose a major global health and economic burden due to their high prevalence and mortality^1,2^. CVD is more prevalent in patients with T2D than in those without, and it represents a leading cause of mortality in this population^3^. Elucidating shared mechanisms is critical for advancing prevention and therapeutic strategies. Branched-chain amino acid (BCAA) metabolism has emerged as a potential shared mechanism^4–6^. Prospective cohort studies have linked elevated circulating levels of BCAAs—isoleucine, leucine, and valine—with increased risk of future onset of both T2D and CVD^4,7,8^. However, some genetic studies suggest that elevated BCAA levels may merely reflect obesity and dyslipidemia, which are traditional risk factors for T2D and CVD^9–11^. Whether BCAA metabolism plays a role in these diseases independent of traditional risk factors remains unclear.

Human genetics has proven to be a powerful approach for identifying and validating therapeutic targets. Drugs with genetic support are more than twice as likely to be approved as those without^12,13^. Recent advances in the field, driven by large-scale genome-wide association studies (GWAS) and increasingly sophisticated post-GWAS methods, have made it possible to disentangle complex biological mechanisms underlying human traits^14–20^. Genomic structural equation modeling (Genomic SEM) is a recently developed framework that enables modeling of shared genetic architecture across traits using GWAS summary statistics^19^. Within this framework, multivariate GWAS can be conducted to identify genetic associations with a latent factor shared across traits, a method referred to as common factor GWAS^19^. Genomic SEM also enables GWAS-by-subtraction, which isolates trait-specific genetic associations by removing effects shared with related traits^20^. These methods have substantially improved our ability to resolve the genetic architecture of complex traits^21–24^.

In this study, we leveraged Genomic SEM to elucidate the role of BCAA metabolism in T2D and CVD independently of traditional risk factors. We first performed a common factor GWAS to identify genetic associations with a latent factor (F_BCAA-Com_) representing shared genetic variance across circulating BCAA levels, using data from individuals of European and East Asian populations.

This analysis incorporated the largest GWAS of circulating BCAA levels in East Asian population to date, comprising up to 42,826 participants from BioBank Japan (BBJ). Next, we examined whether F_BCAA-Com_ associations colocalized with those for body mass index (BMI) and circulating lipid levels. The observed colocalizations at several loci motivated a GWAS-by-subtraction analysis, yielding a latent factor (F_BCAA-Sub_) that captures genetic influences on BCAA levels independently of BMI and lipid levels. Finally, we evaluated the clinical relevance of F_BCAA-Sub_ through association analyses with blood chemistry measurements and the onset of T2D and CVD, aiming to provide human genetic insight into the role of BCAA metabolism beyond traditional risk factors.

## Results

### F_BCAA-Com_ was associated with loci near BCAA metabolism genes

We first investigated genetic correlations among circulating BCAA levels using bivariate linkage disequilibrium score regression (LDSC)^15,16^. GWAS summary statistics were obtained from the UK Biobank (UKB) where BCAA levels were measured using the Nightingale Health nuclear magnetic resonance (NMR) platform (up to n = 115,079; Supplementary Table 1)^25^. Circulating BCAA levels exhibited very high genetic correlations with one another, with the weakest correlation being 0.93 between leucine and valine (Fig. 1a and Supplementary Table 2). To capture the shared genetic structure, we constructed a common factor model in which a single latent factor, referred to as F_BCAA-Com_, influences the levels of isoleucine, leucine, and valine (Fig. 1c). This model demonstrated a good fit to the data (Comparative Fit Index (CFI) = 1.0; standardized root mean square residual (SRMR) = 0.03; Supplementary Table 3; Methods). Consequently, we conducted a common factor GWAS to investigate genetic associations with the F_BCAA-Com_. The genomic inflation factor (λ_GC_ = 1.09) and LDSC intercept (0.981 ± 0.009) indicated no substantial bias (Supplementary Fig. 1). A total of 12 genome-wide significant loci were identified (*P* < 5.0 × 10^−8^; Fig. 2a, Supplementary Table 4), all of which have previously been reported to be associated with circulating BCAA levels. Of the 12 loci, two lead variants, rs1440581 (*PPM1K*) and rs4801775 (*BCAT2*), were located near genes directly involved in BCAA catabolism. PPM1K is a phosphatase that promotes BCAA oxidation, the second step in BCAA catabolism^5^. BCAT2 catalyzes the first step in which BCAAs and alpha-ketoglutarate are converted into branched-chain alpha-keto acids (BCKAs) and glutamate^26^. We also observed lead variants adjacent to genes related to glutamate metabolism: rs7302925 (*SPRYD4*-*GLS2*), rs10211524 (*SLC1A4*), and rs2238732 (*PRODH*). GLS2 is a glutaminase that catalyzes glutamine into glutamate^27^. SLC1A4 is a transporter, and PRODH is a dehydrogenase, both involved in the catabolism of proline into glutamate in mitochondria^28,29^. The remaining lead variants included rs1260326 (*GCKR*) and rs1128249 (*GRB14*-*COBLL1*), both of which have been previously reported to be associated with circulating lipid levels and BMI^30,31^.

**Fig. 1.**
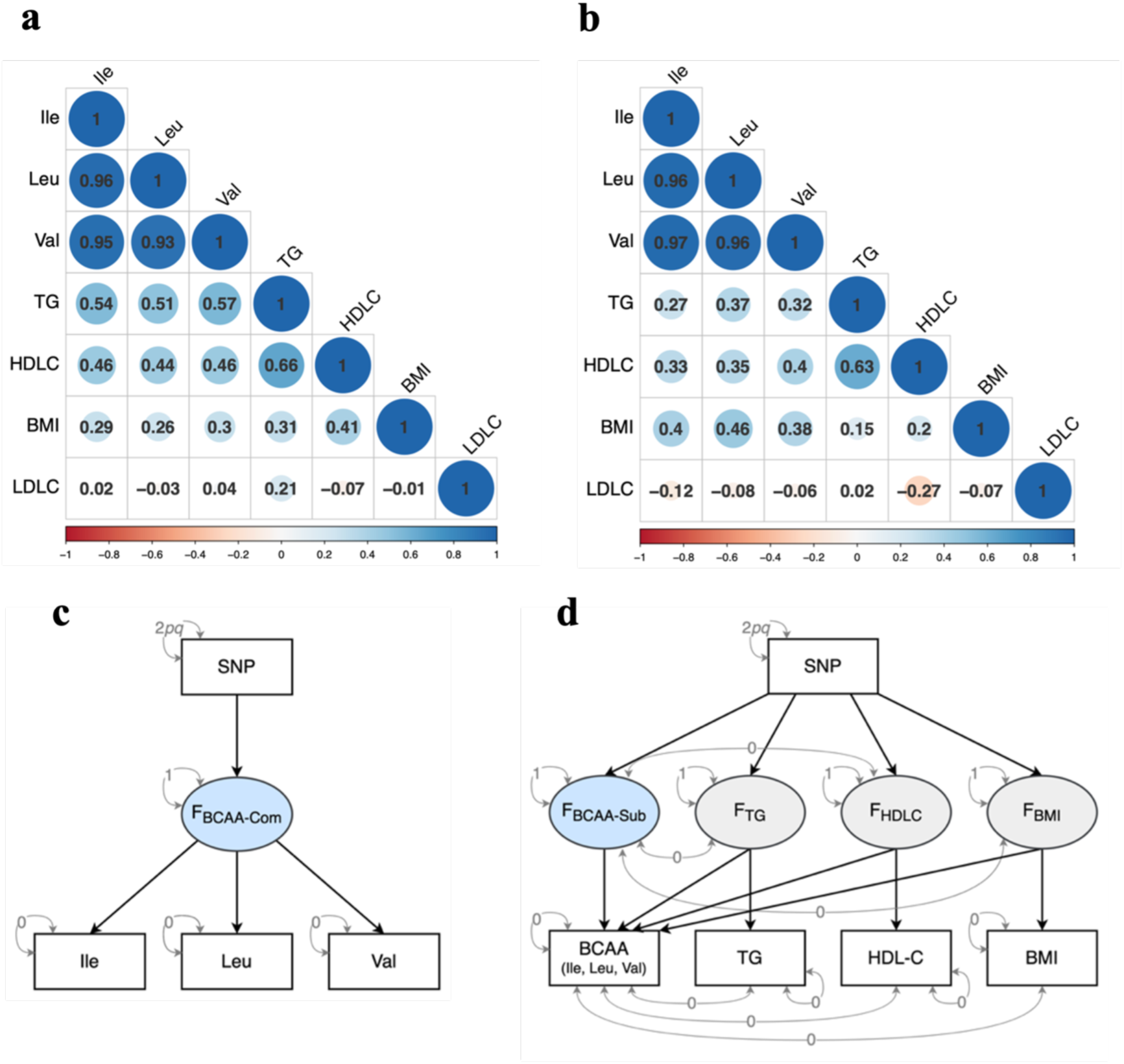
Estimated genetic correlations and Genomic SEM models. **a** and **b**, Estimated genetic correlations among circulating BCAA levels, circulating cholesterol levels, and BMI; **a** in European population and **b** in East Asian population. Shades of blue and red indicate positive and negatic correlations, respectively. **c** and **d**, Path diagram modeling multivariable genetic architecture. The squares represent observed variables based on GWAS summary statistics and the circules represent latent factors. The single-headed arrows are regression relations and the double-headed arrows are variances. The variances of latent factors are fixed to 1. The variance of SNP is fixed to the value of 2*pq* (*p* = reference allele frequency, *q* = alternative allele frequency). The residual variances of observed variables except for SNP are fixed to 0. **c**, The path diagram used for common factor GWAS. **d**, The path diagram used for GWAS-by-subtraction. The covariances between the shared latent factor among BCAAs and the other latent factors and between measurements of BCAAs and the other measurements are fixed to 0.

**Fig. 2.**
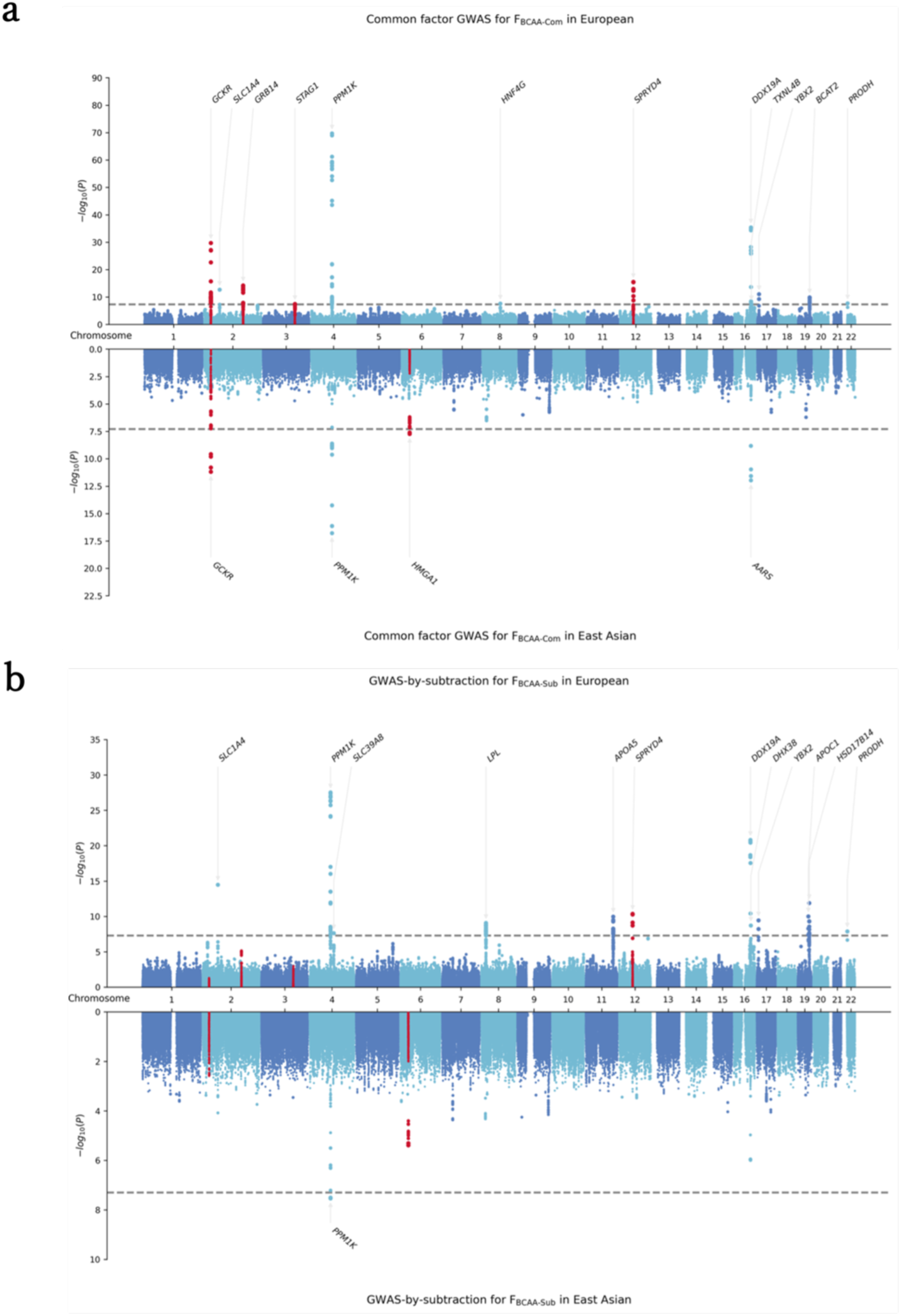
Miami plots of multivariate GWAS for latent factors sharedy by BCAAs. **a**, The Miami plots for F_BCAA-Com_. **b**, The Miami plots for F_BCAA-Sub_. For both plots, the upper panel represents European population, while the bottom panel represents East Asian population in each plot. The grey dashed line indicates the genome-wide significant threshold (*P* < 5.0 × 10^−8^). The red color indicates loci colocalized with any of the circulating cholesterol levels and/or BMI among the genome-wide significant loci.

We next examined whether the shared genetic architecture of BCAAs is also observed in non-European populations. We quantified circulating BCAA levels in 42,843 participants from BBJ using the same platform as the UKB and conducted GWASs (Supplementary Table 1 and Supplementary Fig. 2; Methods). Subsequently, a fixed-effects meta-analysis was performed with GWAS summary statistics from the Tohoku Medical Megabank (TMM) Project, a Japanese project separate from the BBJ (up to n = 7,842; Supplementary Table 1)^32^. Consistent with results from the UKB, genetic correlation analysis revealed remarkably high correlations among BCAAs, with correlation values exceeding 0.95 (Fig. 1b and Supplementary Table 2). The common factor model demonstrated a good fit to the data (CFI = 1.0, SRMR = 0.02; Supplementary Table 3). We then conducted a common factor GWAS, observed no substantial bias (λ_GC_ = 1.08 and LDSC intercept = 0.989 ± 0.009; Supplementary Fig. 1), and identified four genome-wide significant loci (*P* < 5.0 × 10^−8^; Fig. 2b, Supplementary Table 4). The lead variants at the three loci were either identical to or in strong LD with those identified in the above UKB analysis. One such variant was rs1260326 at the *GCKR* locus. At the *PPM1K* and *DDX19B*-*DDX19A* loci, the lead variants were rs7660693 and rs12149660, which are in strong LD with rs1440581 (1000 Genomes Project: EAS R^2^ = 0.88, EUR R^2^ = 0.87) and rs12325419 (EAS R^2^ = 0.85, EUR R^2^ = 0.98), respectively. The remaining variant at the *HGMA1* locus, rs10947487, has not been associated with circulating BCAA levels but is known to be associated with circulating triglyceride (TG) levels and BMI in the East Asian population^33^. Taken together, we identified 13 genome-wide significant loci for the F_BCAA-Com_.

### Some F_BCAA-Com_ loci colocalized with those for lipids and BMI

The common factor GWAS identified loci known to be associated with circulating lipid levels and BMI, suggesting shared genetic associations between BCAA metabolism and these traits. We used HyPrColoc for multi-trait colocalization analysis to assess colocalization at each F_BCAA-Com_ locus^17^. For this analysis in the European population, we used GWAS summary statistics from the Global Lipids Genetics Consortium (GLGC) for circulating TG, high-density lipoprotein cholesterol (HDL-C), and low-density lipoprotein cholesterol (LDL-C) levels (up to n = 888,227; Supplementary Table 1) and from the GIANT consortium for BMI (n = 322,153)^30,34^. We found evidence of colocalizations at four of the 12 loci with at least one lipid level or with BMI (posterior probability of full colocalization (PPFC) > 0.7; Fig. 3a and Supplementary Table 5; Methods). For example, the *GCKR* locus colocalized between F_BCAA-Com_ and TG (PPFC = 0.72), with the PPFC fully explained by rs1260326. At the *GRB14*-*COBLL1* locus, we observed colocalization with multiple traits: TG, HDL-C, and BMI (PPFC = 0.72 of which 97.3% was explained by rs1128249). Colocalization analysis showed no strong evidence at the remaining eight loci, including those near genes involved in BCAA catabolism such as *PPM1K* and *BCAT2*.

**Fig. 3.**
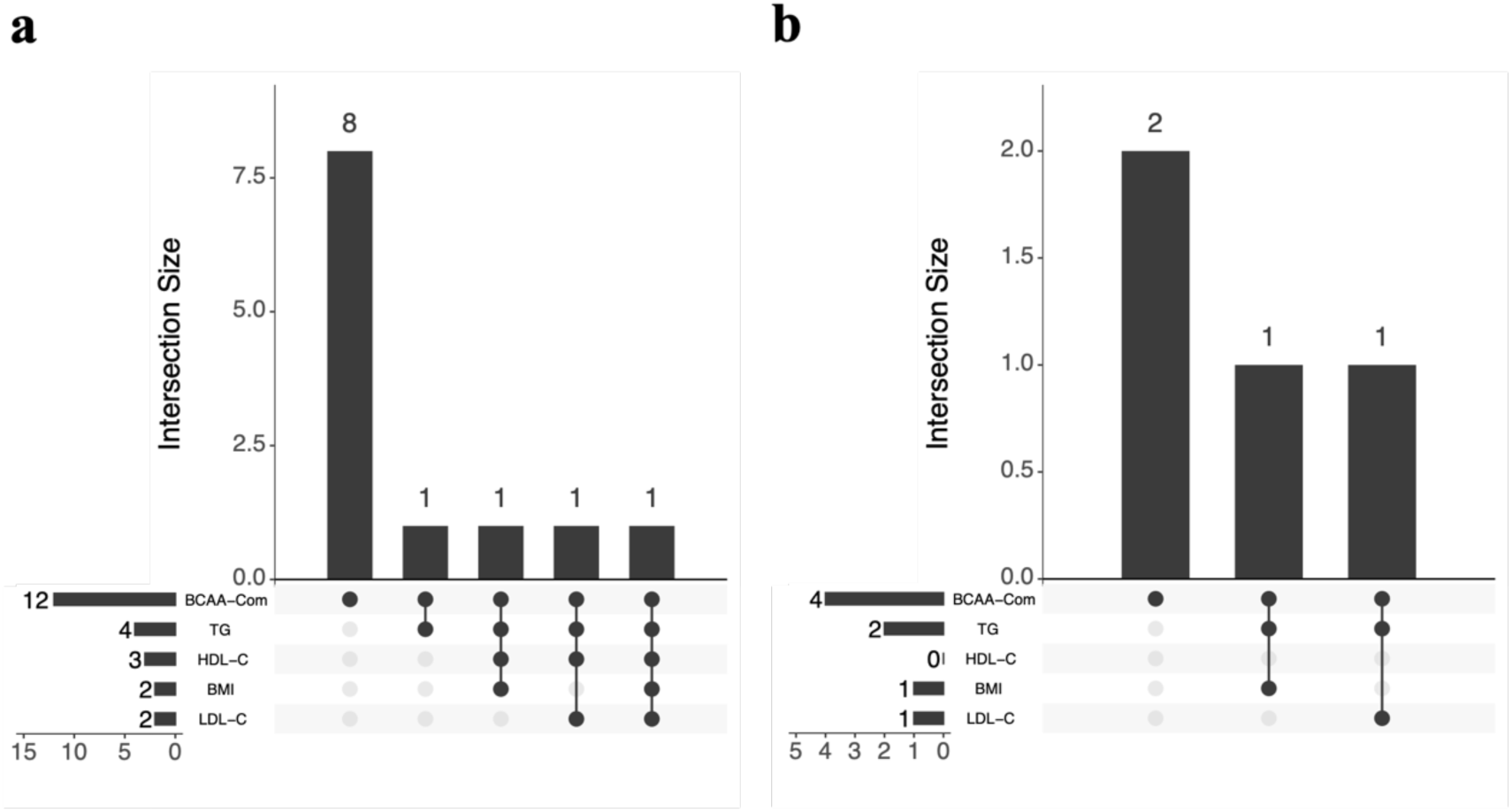
Colocalized loci between the F_BCAA-Com_ and circulating cholesterol levels and BMI. **a** and **b**, The upset plots represent the colocalized loci between F_BCAA-Com_ and circulating TG levels, HDL-C levels, LDL-C levels, and BMI, restricted to loci significantly associated with F_BCAA-Com_, **a** in European population, and **b** in East Asian population. The left bars indicate the total number of colocalized loci with the F_BCAA-Com_ for each trait. The intersection size indicates the number of colocalized loci shared among traits represented as points connected with vertical lines.

To validate shared genetic associations between F_BCAA-Com_ and both circulating lipid levels and BMI in the East Asian population, we conducted a multi-trait colocalization analysis using GWAS summary statistics from the Korean Genome and Epidemiology Study (KoGES: up to n = 72,297; Supplementary Table 1)^33^. Of the four F_BCAA-Com_ loci, we found evidence of colocalization at two loci with at least one lipid level or BMI (Fig. 3b and Supplementary Table 5). We observed colocalization again at the *GCKR* locus between F_BCAA-Com_ and TG (PPFC = 0.83), with the PPFC fully explained by rs1260326. As expected, the *HMGA*1 locus was genetically shared across F_BCAA-Com_, TG, and BMI (PPFC = 0.96 of which 65.2% was explained by rs10947487). The remaining *PPM1K* and *DDX19B*-*DDX19A* loci showed no strong evidence of colocalization with any lipid levels or BMI. Our findings indicate that BCAA metabolism is genetically influenced by latent factors whose genetic architectures are either shared with circulating lipid levels and BMI or independent of them, motivating further analyses to disentangle the latter.

### F_BCAA-Sub_ influenced circulating BCAA levels independently of traditional risk factors

We used GWAS-by-subtraction to investigate a genetic architecture shared by BCAAs, independent of traditional risk factors. We used the same GWAS summary statistics as in the colocalization analysis, without LDL-C due to the weak and nonsignificant genetic correlations with each BCAA (Fig. 1a, b and Supplementary Table 2). To explore a model that fits the data well, we compared three models with different configurations for TG, HDL-C, and BMI (Fig. 1d and Supplementary Fig. 3; Methods). The model with the lowest Akaike Information Criterion (AIC) value comprises four latent factors: one influencing only BCAAs, referred to as F_BCAA-Sub_, and three others that influence TG, HDL-C, and BMI, respectively, in addition to BCAAs (Fig. 1d and Supplementary Table 3). This model also demonstrated a good fit in absolute indices (European: CFI = 1.0, SRMR = 0.02; East Asian: CFI = 1.0, SRMR = 0.01; Supplementary Table 3). Thus, we used this model in subsequent analyses.

GWAS-by-subtraction identified 12 genome-wide significant loci (*P* < 5.0 × 10^−8^) without substantial bias in the European population (λ_GC_ = 1.05 and LDSC intercept = 0.969 ± 0.007; Fig. 2b, Supplementary Fig. 1 and Supplementary Table 6). Notably, among the four loci where we identified colocalization with the lipid levels and/or BMI, three did not reach the genome-wide significance, including the *GCKR* locus, substantiating the utility of GWAS-by-subtraction.^41,42^ For East Asian population, GWAS-by-subtraction identified one genome-wide significant locus (*P* < 5.0 × 10^−8^) at *PPM1K* without substantial bias (λ_GC_ = 1.05 and LDSC intercept = 0.943 ± 0.008; Supplementary Fig. 1). The *GCKR* locus did not reach genome-wide significance. To assess whether the genetic architecture of the F_BCAA-Sub_ is shared across European and East Asian populations, we compared the effect sizes of lead variants at significant loci. This analysis revealed fully concordant directions of effect and a strong correlation (Pearson’s correlation coefficient = 0.89, *P* = 6.4 × 10^−4^; Supplementary Fig. 4). We further conducted a cross-population genetic correlation analysis using Popcorn and observed a substantial genetic correlation (*ρ*_gi_ = 0.65, standard error = 0.23)^35^. These findings support a shared genetic architecture underlying the F_BCAA-Sub_ across populations.

Next, we built polygenic scores (PGS) for each latent factor using summary statistics from common factor GWAS and GWAS-by-subtraction. To improve cross-population polygenic prediction, we used PRS-CSx, which integrates summary statistics from multiple populations (Supplementary Fig. 5; Methods)^18^. We first assessed the PGS characteristics using 4,760 BBJ participants who had measurements for circulating BCAA levels and were not used for PGS construction (Methods). The F_BCAA-Com_ PGS showed positive associations with circulating TG and HDL-C levels and BMI in linear regression analyses adjusted for age, age-squared, sex, and the first ten genetic principal components (TG: β = 0.09, 95% confidence interval (CI) = 0.06–0.12; HDL-C: β = 0.05, 95% CI = 0.02–0.08; BMI: β = 0.06, 95% CI = 0.03–0.10; Fig. 4a and Supplementary Table 7). In contrast, the effect sizes for the F_BCAA-Sub_ PGS remained consistently close to zero for TG, HDL-C, and BMI, with confidence intervals including zero (TG: β = 0.01, 95% CI = −0.02*‒*0.03; HDL-C: β = −0.01, 95% CI = −0.04–0.02; BMI: β = 0.02, 95% CI = −0.02–0.05). These results are consistent with the GWAS-by-subtraction model, which assumes that the F_BCAA-Sub_ is independent of lipid- and BMI-related latent factors. We next tested associations between the F_BCAA-Sub_ PGS and circulating BCAA levels, and found consistently positive effects in a baseline model adjusted for age, age-squared, sex, the first ten genetic principal components, and metabolite measurement-related covariates (Ile: β = 0.04, 95% CI = 0.02–0.07; Leu: β = 0.05, 95% CI = 0.02–0.08; Val: β = 0.06, 95% CI = 0.04–0.09; Fig. 4b and Supplementary Table 8; Methods). These associations remained robust after further adjustment for the three additional PGSs corresponding to latent factors associated with TG, HDL-C, and BMI (Ile: β = 0.05, 95% CI = 0.02–0.08; Leu: β = 0.05, 95% CI = 0.03–0.08; Val: β = 0.07, 95% CI = 0.04–0.10).

**Fig. 4.**
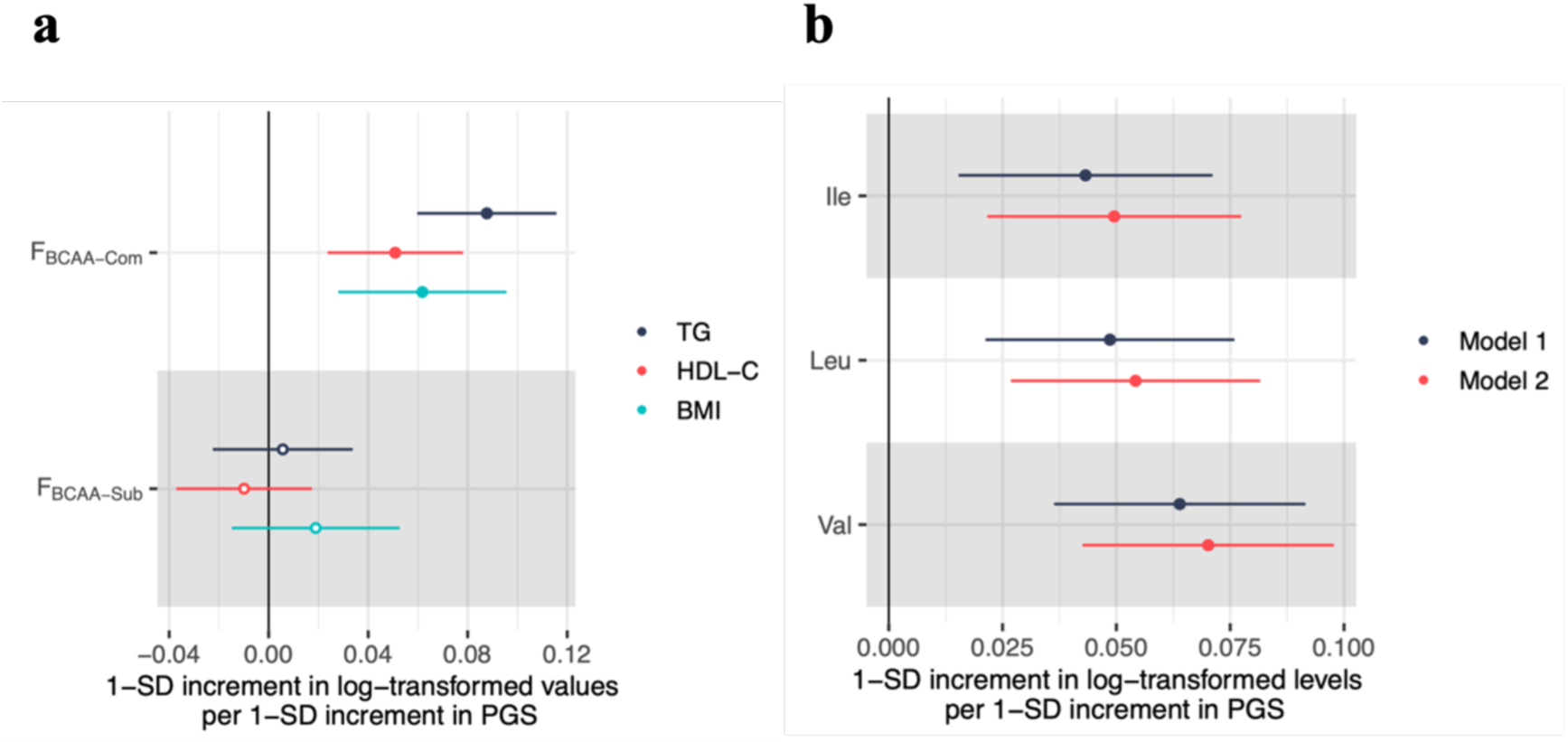
Associations between latent factor PGSs and cholesterols, BMI, and BCAAs. **a**, The forest plot illustrates the associations between a 1-SD increment in PGSs for F_BCAA-Com_ and F_BCAA-Sub_ and a 1-SD increment in the log-transformed values of circulating TG level, HDL-C level, and BMI, represented in black, red, and cyan, respectively. The solid circles denote statistically significant associations (Bonferroni-corrected threshold *P* < 0.05/6). **b**, The forest plot illustrates the associations between a 1-SD increment in F_BCAA-Sub_ PGS and a 1-SD increment in the log-transformed circulating BCAA levels. The black indicate a result of linear regression analysis not adjusted for any PGSs of F_TG-Sub_, F_HDL-C-Sub_, and F_BMI-Sub_ (Model 1), while the red indicate a result adjusted for these latent factors (Model 2). The solid circles denote statistically significant associations (Bonferroni-corrected threshold *P* < 0.05/6). The error bars represent 95% CIs.

This extended model consistently reduced AIC values across all three BCAAs and increased adjusted R^2^ by up to 12.5% relative to the baseline model (ANOVA *P*_Ile_ = 5.4 × 10^−7^; *P*_Leu_ = 1.2 × 10^−5^; *P*_Val_ = 6.5 × 10^−8^; Supplementary Table 9). These results indicate that circulating BCAA levels reflect a composite phenotype shaped by distinct genetic latent factors, including factors independent of or shared with lipid levels and BMI.

The three BCKAs*̶*3-methyl-2-oxovalerate (KMV), 4-methyl-2-oxopentanoate (KIC), and 3-methyl-2-oxobutyrate (KIV)*̶*are known to respond to the modulation of BCAA catabolism without changes in circulating lipid levels or body weight in mice^36–38^. Given this, we hypothesized that the F_BCAA-Sub_ may reflect a genetic factor related to BCAA catabolism and therefore performed genetic correlation analyses between the F_BCAA-Sub_ and circulating BCKA levels. This analysis was restricted to the European population due to the need for robust estimation requiring large sample sizes (Supplementary Table 1; Methods)^23,39,40^. In line with our hypothesis, the F_BCAA-Sub_ exhibited the highest magnitude of the correlation coefficient with all BCKAs, followed by the latent factors for TG, HDL-C, and BMI (F_BCAA-Sub_: *r*_g_ = 0.32–0.33; F_TG-Sub_: *r*_g_ = 0.20–0.21; F_HDL-C-Sub_: *r*_g_ = 0.16; F_BMI-Sub_: *r*_g_ = 0; Supplementary Fig. 6 and Supplementary Table 10). These findings indicate that the F_BCAA-Sub_ preferentially reflects a genetic component related to BCAA catabolism rather than a general cardiometabolic axis.

### F_BCAA-Sub_ was associated with glucose metabolism and organ dysregulation

To better understand the clinical characteristics of F_BCAA-Sub_, we conducted comprehensive association analyses with 18 blood chemistry tests, categorized into six groups: liver function tests, proteins, glucose metabolism, kidney function tests, electrolytes, and enzymes. Using data from 134,176 participants from the BBJ who were not included in PGS construction (Supplementary Fig. 5), we identified five significant associations, in linear regression analyses adjusted for age, age-squared, sex, and the first ten genetic principal components, at a Bonferroni-corrected threshold (*P* < 0.05/18; Fig. 5 and Supplementary Table 11). Positive associations were identified between the F_BCAA-Sub_ PGS and two traits related to glucose metabolism: blood glucose (BG; *β* = 0.02, 95% CI = 0.02–0.03) and hemoglobin A1c (HbA1c; *β* = 0.03, 95% CI = 0.02–0.04). We also identified positive associations with alanine transaminase (ALT; *β* = 0.01, 95% CI = 0.004–0.01) and blood urea nitrogen (BUN; *β* = 0.02, 95% CI = 0.01–0.02). ALT and BUN have been associated with future diabetes risk in previous studies^41,42^. To investigate whether the latent factor is associated with these tests prior to the onset of T2D, we performed regression analyses, restricting the analysis to participants never diagnosed with T2D. Four associations remained significant, except for potassium (*P* < 0.05/18; Supplementary Fig. 7 and Supplementary Table 11). Our findings suggest that the genetic factor influencing BCAA metabolism may contribute to glucose metabolism and organ dysregulation prior to the onset of T2D, independent of circulating lipid levels and BMI.

**Fig. 5.**
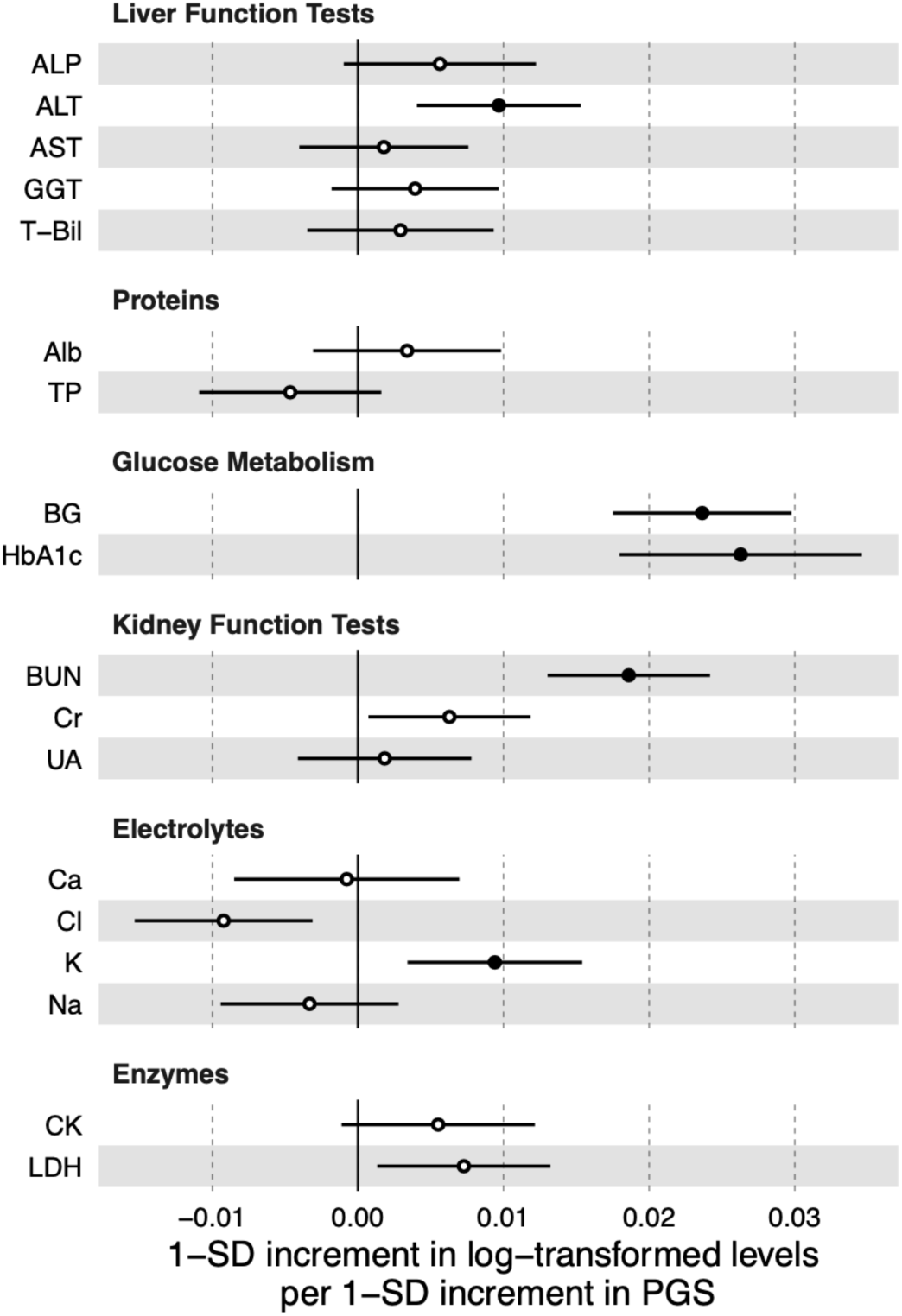
Associations between the F_BCAA-Sub_ and 18 blood chemistry tests. The forest plot illustrates the associations between a 1-SD increment in F_BCAA-Sub_ PGS and a 1-SD increment in log-transformed levels of 18 blood chemistry tests. The error bars represent 95% CIs. The results are from linear regression analyses conducted on all individuals not used in PGS construction. The solid circles denote statistically significant associations (Bonferroni-corrected threshold *P* < 0.05/18). ALP, alkaline phosphatase; ALT, alanine aminotransferase; AST, aspartate aminotransferase; GGT, gamma-glutamyl transferase; T-Bil, total bilirubin; Alb, albumin; TP, total protein; BG, blood glucose; HbA1c, hemoglobin A1c; BUN, blood urea nitrogen; Cr, creatinine; UA, uric acid; Ca, calcium; Cl, chloride; K, potassium; Na, sodium; CK, creatine kinase; LDH, lactate dehydrogenase.

To gain deeper insight into the relationship between F_BCAA-Sub_ and insulin resistance (IR), we examined the genetic correlations between F_BCAA-Sub_ and seven IR-related traits. Fasting insulin (FI), fasting glucose (FG), and homeostasis model assessment of insulin resistance (HOMA-IR) reflect hepatic insulin sensitivity in the fasting state. Two-hour post-challenge glucose (2hGlu), insulin fold change (IFC), and the modified Stumvoll insulin sensitivity index (ISI) represent postprandial IR. Waist-to-hip ratio (WHR) was also included in the analysis. This analysis was restricted to the European population due to the need for robust estimation requiring large sample sizes (Supplementary Table 1)^43–46^. The F_BCAA-Sub_ showed a negative correlation with ISI (*r*_g_ = −0.18; Supplementary Fig. 8 and Supplementary Table 12). The correlation coefficients were positive for the other traits (*r*_g_ = 0.07–0.16). These results consistently indicate a positive correlation between F_BCAA-Sub_ and both fasting and postprandial IR.

### F_BCAA-Sub_ PGS predicted the onset of T2D and CAD

Given the observed associations between F_BCAA-Sub_ and glycemic traits, we investigated whether F_BCAA-Sub_ also plays a role in T2D. Using the same BBJ dataset as in the PGS analysis on blood chemistry tests, we conducted a logistic regression analysis to examine the association between the F_BCAA-Sub_ PGS and the onset of T2D, diagnosed at recruitment or during follow-up. A positive association was identified in a baseline model adjusted for age, age-squared, sex, and the first ten genetic principal components (odds ratio (OR) = 1.07, 95% CI = 1.06–1.09; Fig. 6a and Supplementary Table 13; Methods). This association remained positive after additional adjustment for circulating lipid levels and BMI (OR = 1.06, 95% CI = 1.04–1.09), and was robust to further adjustment for smoking history, drinking history, systolic blood pressure, and diastolic blood pressure (OR = 1.07, 95% CI = 1.04–1.10). Next, we divided participants into deciles based on the F_BCAA-Sub_ PGS and performed a survival analysis to investigate differences in age at diagnosis among the top, middle, and bottom deciles. Across these strata, no notable differences were observed in follow-up duration, age at recruitment, sex, or any risk factors (Supplementary Table 14). The survival analysis revealed that participants in the top decile were diagnosed with T2D at a younger age than those in the bottom decile (log-rank test, *P* = 1.1 × 10^−16^; Fig. 6d). Similar patterns were observed with alternative stratification thresholds of 5% and 20% (Supplementary Fig. 9).

**Fig. 6.**
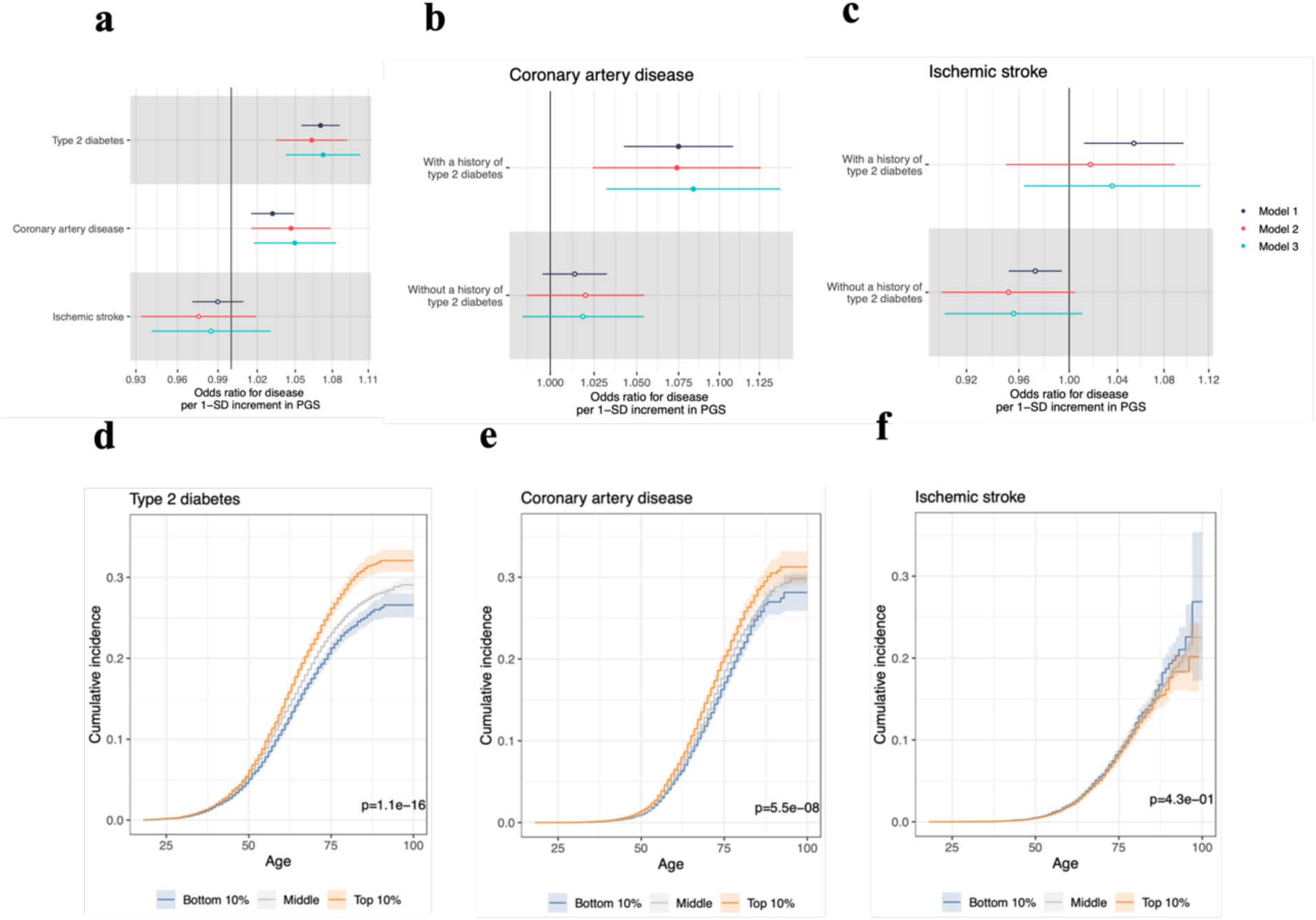
**Associations between the F_BCAA-Sub_ and cardiometabolic diseases. a-c**, The forest plots illustrate the associations between a 1-SD increment in F_BCAA-Sub_ PGS and the odds ratio for cardiometabolic diseases. The error bars represent 95% CIs. The results of linear regression analyses using the baseline model in black (Model 1), the model adjusted for circulating cholesterol levels and BMI in red (Model 2), and the model additionally adjusted for cardiometabolic risk factors in cyan (Model 3). **a**, Evaluation of the onset of type 2 diabetes (T2D), coronary artery disease (CAD), and ischemic stroke, respectively. The solid circles and open circles denote statistically significant and non-significant associations, respectively (Bonferroni-corrected threshold *P* = 0.05/9). **b** and **c**, Evaluations of comorbidity of T2D with CAD and ischemic stroke. The solid circles and open circles denote statistically significant and non-significant associations, respectively (Bonferroni-corrected threshold *P* = 0.05/6). **d-f**, The cumulative incidence curves for the onset of each disease (**d**, T2D; **e**, CAD; **f**, ischemic stroke) stratified by PGS strata: bottom 10% in blue, middle in gray, and top 10% in orange. The shared area represent 95% CIs. The *P* values from the log-rank tests are displayed in the bottom-right corner.

We extended our investigation to include coronary artery disease (CAD) and ischemic stroke, two leading causes of CVD mortality^52^. Using three covariate adjustment models, as in the T2D analyses, we identified a consistent positive association between the F_BCAA-Sub_ PGS and the onset of CAD (OR = 1.03–1.05, 95% CI = 1.02–1.08; Fig. 6a and Supplementary Table 13). Survival analyses across different stratification thresholds consistently showed earlier CAD diagnosis among participants in the top decile compared with those in the bottom decile (log-rank test, *P* = 5.5 × 10^−8^; Fig. 6e and Supplementary Fig. 10), while no significant associations were observed between the F_BCAA-Sub_ and the onset of ischemic stroke (OR = 0.98–0.99, 95% CI = 0.93–1.03; Fig. 6a, 6f and Supplementary Fig. 11). Subgroup analyses by ischemic stroke subtype—atherothrombotic, cardioembolic, and lacunar—also showed no significant associations (Supplementary Table 15). Lastly, we investigated the association between the F_BCAA-Sub_ PGS and the comorbidity of T2D and CVD. Logistic regression analyses were performed separately for participants with and without a history of T2D to assess the association with CVD onset. Notably, the F_BCAA-Sub_ PGS was positively associated with CAD onset in participants with T2D (OR = 1.07–1.08, 95% CI = 1.02–1.14), with no clear association in those without T2D (Fig. 6b, 6c and Supplementary Table 16). These findings demonstrate that the F_BCAA-Sub_ PGS predicts both T2D and CAD, supporting a shared genetic architecture underlying their comorbidity.

## Discussion

T2D and CVD remain among the leading global health burdens, and understanding their shared biological mechanisms is critical for prevention and therapeutic development. BCAA metabolism has been implicated in these diseases, but whether it plays an independent role beyond traditional risk factors remains unclear. By leveraging Genomic SEM of large-scale GWAS summary statistics, we identified a latent factor (F_BCAA-Sub_) that captures genetic influence on circulating BCAA levels independently of circulating lipid levels and BMI. We then found that this factor is associated with glucose metabolism, T2D, and CAD. These associations remained significant after adjustment for other traditional risk factors, including smoking, drinking, and blood pressure. Our study provides the first genetic insight in humans that BCAA metabolism is involved in T2D and CAD beyond traditional risk factors, highlighting its clinical relevance and therapeutic potential.

This study investigated genetic architecture underlying BCAA metabolism in humans, with a focus on its potential relevance to T2D and CVD. Although elevated circulating BCAA levels have been linked to disease risk, it remains unclear whether these associations are independent of traditional risk factors such as obesity and dyslipidemia^7,8^. To address this, we identified the genetic latent factor, F_BCAA-Sub_, which is associated with circulating BCAA levels independently of BMI and circulating lipid levels. This finding points to the presence of an endogenous mechanism regulating BCAA metabolism, not explained by traditional risk factors. The F_BCAA-Sub_ appears to reflect genetic influence on BCAA catabolism, as supported by its genetic correlations with BCKAs, intermediate metabolites of BCAA catabolism, and the proximity of associated loci to *PPM1K*, a gene known to play a critical role in the rate-limiting step of BCAA catabolism^5^. Furthermore, the F_BCAA-Sub_ was positively associated with blood glucose, HbA1c, and ALT, implying potential alterations in glucose metabolism and liver function. These findings are consistent with rodent studies demonstrating that modulating BCAA catabolism alters BCKA levels, IR, and hepatic oxidative stress, without affecting body weight or circulating lipid levels^36–38,47^.

Notably, the F_BCAA-Sub_ was associated with increased risk of both T2D and CAD, even after adjusting for circulating lipid levels, BMI, smoking, drinking, and blood pressure. These findings underscore the potential contribution of BCAA metabolism to these diseases beyond traditional risk factors. We also found that the association between the F_BCAA-Sub_ and CAD was stronger in individuals with T2D than in those without. This is consistent with previous epidemiological evidence pointing to a shared pathophysiological mechanism involving BCAA metabolism in both T2D and CVD^4^. Our genetic findings expand on this evidence by demonstrating that the involvement of BCAA metabolism remains robust even after accounting for potential confounding by traditional risk factors. Although associations with blood glucose and HbA1c suggest glucose metabolism as a shared mechanism, the molecular mechanisms underlying these relationships remain incompletely understood, highlighting the need for further functional studies.

Our findings provide several insights relevant to the clinical translation of BCAA metabolism as a therapeutic target. First, the independence of the F_BCAA-Sub_ from traditional risk factors suggests that BCAA metabolism may represent a novel therapeutic target. A short-term, randomized clinical trial has shown that pharmacological enhancement of BCAA catabolism improves insulin sensitivity in patients with T2D^48^. While further clinical trials are warranted to evaluate its efficacy, our genetic findings provide additional rationale for future clinical trials^12^. Second, individuals with higher F_BCAA-Sub_ PGS but no prior diagnosis of T2D exhibited elevated blood glucose and HbA1c levels. This observation highlights the value of further investigating BCAA metabolism as a therapeutic target to delay or prevent disease onset in at-risk individuals. Third, our cross-population analyses revealed shared genetic effects across European and East Asian populations, supporting the potential applicability of targeting BCAA metabolism as a therapeutic strategy across diverse populations. Collectively, these genetic findings pave the way for the clinical translation of BCAA metabolism.

There are limitations to the present study. First, GWAS-by-subtraction assumes no correlation between the F_BCAA-Sub_ and the other latent factors. This assumption may be overly simplistic for accurately capturing the complexity of BCAA metabolism. Nevertheless, the model exhibited good fit and yielded results consistent with prior studies. Second, this study focused solely on genetic architecture as an intrinsic factor. BCAA restriction has been demonstrated to improve IR in both rodent and human studies^49,50^. Elucidating the interaction between genetic architecture and dietary intake may yield deeper insights into BCAA metabolism and guide the development of more effective therapeutic strategies.

Despite these limitations, this study identified the genetic factor, F_BCAA-Sub_, that influences circulating BCAA levels independently of traditional risk factors in humans. This factor predicts future risk of T2D and CAD. Our study provides genetic evidence that BCAA metabolism plays a role in T2D and CAD beyond traditional risk factors, highlighting its clinical relevance and therapeutic potential.

## Methods

### GWAS summary statistics and the BBJ Project

GWAS summary statistics from large-scale studies were used for the genetic analyses described below. Detailed information about each GWAS summary statistics is provided in Supplementary Table 1.

The BBJ Project is a hospital-based study in Japan^51^. This project recruited approximately 200,000 participants who were diagnosed with at least one of the 47 target diseases from 2003 to 2008, as the first cohort. This project collected DNA, serum samples, and clinical data at baseline from 66 hospitals across Japan and continued to collect serum samples and clinical data annually until 2013. This project was approved by the ethics committee of the Institute of Medical Science, the University of Tokyo (approval number: 2023-77-0118). All participants provided informed consent.

### Metabolite measurements

Metabolomic analysis was conducted on serum samples collected from BBJ participants^51^. All serum samples were initially stored at −80°C at cooperating hospitals. After several samples were stored, samples were delivered to the BBJ and stored at −150 °C in the BBJ serum bank. The serum samples were shipped to Nightingale Health laboratories in Tokyo in 96-well plates on dry ice in batches of approximately 10,000 samples. Samples were stored in a freezer at −80 °C at Nightingale Health laboratories after arrival from BBJ laboratory. Before preparation, frozen samples were slowly thawed at +4 °C overnight, and then mixed gently and centrifuged (3 min, 3400 × g, +4 °C) to remove possible precipitate. Aliquots of each sample were transferred into 3-mm outer-diameter NMR tubes and mixed in 1:1 ratio with a phosphate buffer (75 mM Na2HPO4 in 80%/20% H2O/D2O, pH 7.4, including also 0.08% sodium 3-(trimethylsilyl) propionate-2,2,3,3-d4 and 0.04% sodium azide) automatically with an automated liquid handler (PerkinElmer Janus Automated Workstation)^52^. This platform quantified a total of 250 metabolic traits, including BCAAs. All serum samples used in this study were analyzed independently of the target diseases.

### Genotyping, quality control, and imputation

Samples in the BBJ were genotyped using the Illumina HumanOmniExpressExome BeadChip or a combination of the Illumina HumanOmniExpress and HumanExome BeadChip. Detailed quality control of participants and genotypes have been described in the previous study^53^. Pre-phasing was performed using Eagle (v.2.4.1) without an external reference panel^54^. Imputation was performed using Minimac4 (v.1.0.2) with the JEWEL3k reference panel (3,256 Japanese whole-genome sequence data and 2,504 1000 Genomes Project Phase 3, version 5)^55,56^. The coordinates were aligned to the human genome build hg19.

### GWAS

We performed quality control and divided the dataset to conduct GWASs on circulating BCAA levels in the BBJ and to perform subsequent analyses (Supplementary Fig. 2). As part of the quality control process, we first selected samples from participants aged 18 or older, with available imputed genotype data and no missing values for age or sex. Next, we retained samples that passed measurement quality control by Nightingale Health and included only the earliest sample from each participant. Finally, we selected samples from unrelated participants. A total of 47,603 samples passed the quality control steps. Ninety percent of these samples (n = 42,843) were randomly selected for GWAS, and the remaining 10% (n = 4,760) were used as an independent test dataset to evaluate PGS performance.

Before conducting the GWAS, BCAA levels were log-transformed, and values exceeding 4 standard deviations from the mean were treated as missing. The values were adjusted for age, age-squared, sex, sampling month, NMR shipping batch, processing duration, spectrometer, and the first ten genetic principal components. The residuals were subsequently normalized using an inverse-normal transformation. We then conducted the GWAS using BOLT-LMM (v.2.4.1)^57^, a linear mixed model-based method that accounts for population stratification, analyzing the imputed autosomal variants with Rsq ≥ 0.3 and minor allele frequency (MAF) > 0.01. In total, more than 14.9 million variants were included in the analysis.

### Meta-analysis

Meta-analyses were conducted using a fixed-effects inverse-variance meta-analysis implemented in METAL (v.2011-03-25)^58^. For circulating levels of each BCAA (isoleucine, leucine, and valine), we performed meta-analyses on GWAS summary statistics from the present study and the TMM Project, a Japanese project separate from the BBJ. For circulating levels of each BCKA (KMV, KIC, and KIV), we performed meta-analyses on GWAS summary statistics from three European cohorts. Details of each GWAS summary statistics are provided in Supplementary Table 1.

### LDSC and Popcorn

We used LDSC (v.1.0.1) to examine bias from population stratification or cryptic relatedness and to estimate genetic correlations^15,16^. We used Popcorn (v.1.0.0) to estimate cross-population genetic correlations between European and East Asian populations^35^. We performed the analyses with population-matched LD scores provided by the developers, restricting the analysis to HapMap3 variants with MAF > 0.01. Details of the GWAS summary statistics used in the analysis are provided in Supplementary Table 1. The direction of effect sizes for circulating HDL-C levels was reversed to align with those for circulating TG levels and BMI, reflecting their role as indicators of cardiometabolic risk factors.

### Common factor GWAS

Genomic SEM (v.0.0.5) was used to investigate genetic associations with a latent factor that commonly influences circulating levels of isoleucine, leucine, and valine^19^. We constructed a model in which a single latent factor (F_BCAA-Com_) influences these levels (Fig. 1c). The models were constructed separately for European and East Asian populations, using GWAS summary statistics from the UK Biobank (up to n = 115,079; Supplementary Table 1) and from the meta-analysis described above, respectively. Model fit was evaluated using the CFI, SRMR, and AIC. For CFI and SRMR, values greater than 0.95 and less than 0.05, respectively, were considered indicative of a good fit^19^. AIC is a relative index, with lower values indicating a better fit.

HapMap3 variants with a MAF > 0.01, as reported in the summary statistics of each study, were used to investigate genetic associations with the latent factor. Genome-wide significant loci were determined by iteratively extending 500-kb flanking regions around genome-wide significant variants (*P* < 5.0 × 10^−8^) until no such variants were included. The variant with the lowest p-value within each locus was defined as the lead variant. Association test results were visualized in a Miami plot using GWASLab (v.3.4.45)^59^. The effective sample size for the latent factor was calculated using the previously described formula for the following analyses^20,60^.

### Colocalization analysis

Multi-trait colocalization analysis was conducted using HyPrColoc (v.0.0.2), which can identify colocalization across multiple traits simultaneously using GWAS summary statistics^17^. We performed a colocalization analysis at each genome-wide significant locus in the common factor GWAS to investigate colocalizations with BMI and circulating levels of TG, HDL-C, and LDL-C. The analyses were performed separately for European and East Asian populations. All GWAS summary statistics for lipid levels and BMI were derived from biobank-scale studies (Supplementary Table 1). HyPrColoc assumes summary statistics are derived from studies with no overlapping individuals. To minimize violating this assumption, we used summary statistics from cohorts different from those in the common factor GWAS. However, the developers have demonstrated that colocalizations can still be correctly detected even if this assumption is violated. We performed colocalization analysis using the variant specific configuration priors with the regional threshold (*P_R_*\* = 0.7) and alignment threshold (*P_A_*\* = 0.7). In accordance with the recommendation provided by the developers, we reported colocalizations whose PPFC exceeded 0.7 (*P_R_P_A_* > 0.7)^17^.

### GWAS-by-subtraction

GWAS-by-subtraction was conducted using Genomic SEM (v.0.0.5)^19,20^. We constructed three models constrained to have no correlations between the latent factor for circulating BCAA levels (F_BCAA-Sub_) and the latent factor(s) for circulating TG levels, HDL-C levels, and BMI (Supplementary Fig. 3). Circulating LDL-C levels were excluded from these models due to the weak and nonsignificant genetic correlations with circulating BCAA levels (|*r*_g_| < 0.1 and *P* > 0.05 for all pairs, except for Ile in East Asian population: *r*_g_ = −0.12, *P* = 0.32; Supplementary Table 2). One model consists of the F_BCAA-Sub_ and three latent factors that influence TG, HDL-C, and BMI, respectively, in addition to BCAAs (Model 1). The other two models are similar but differ in that one includes a latent factor shared by TG and HDL-C (Model 2), and the other includes a latent factor shared by both lipid levels and BMI (Model 3). These models were constructed separately for European and East Asian populations, based on GWAS summary statistics for lipid levels and BMI used in the colocalization analysis, in addition to those for BCAAs used in the common factor GWAS. The direction of effect sizes for circulating HDL-C levels was reversed to align with those for circulating TG levels and BMI, reflecting their role as indicators of cardiometabolic risk factors. Model fit was evaluated using the same indices as in the common factor GWAS. Model 1, consisting of four latent factors, showed the best fit. Thus, this model was used for the following analyses. HapMap3 variants with a MAF > 0.01, as reported in the summary statistics of each study, were used to investigate genetic associations with each latent factor. The same process used for the common factor GWAS was applied to derive genome-wide significant loci, lead variants, and estimated sample sizes.

### PGS

For each latent factor modeled in the common factor GWAS and GWAS-by-subtraction, PGS weights were estimated using PRS-CSx (v.1.0.0) that improves cross-population polygenic prediction by jointly modeling GWAS summary statistics from multiple populations (Supplementary Fig. 5)^18^. We used the ’auto’ option, which automatically learns the global shrinkage parameter from the discovery summary statistics. We also used the ’meta’ option, which integrates population-specific posterior variant effects using inverse-variance-weighted meta-analysis. Combining these options enables the generation of cross-population PGS without the need for an additional validation dataset. We used population-matched LD reference panels provided by the developer.

Evaluation of PGSs was performed using the BBJ participants described above (n = 4,760), who were independent of the PGS construction. For each participant, PGS was calculated as the sum of the dosages multiplied by the PGS weight for each variant. We evaluated the F_BCAA-Com_ and F_BCAA-Sub_ PGS for predicting circulating TG levels, HDL-C levels, and BMI using a linear regression model adjusted for age, age-squared, sex, and the first ten genetic principal components. We also evaluated the F_BCAA-Sub_ PGS for predicting circulating BCAA levels using a linear regression model adjusted for age, age-squared, sex, sampling month (as a proxy for seasonality), NMR shipping batch, processing duration, spectrometer, and the first ten genetic principal components. This evaluation showed that cross-population weights predicted all BCAA levels better than population-specific weights (Supplementary Table 17). Therefore, we adopted cross-population weights to calculate PGS for all subsequent PGS analyses.

### Statistical analysis

Regression analyses were conducted to investigate the relationships between the F_BCAA-Sub_ and both blood chemistry tests and cardiometabolic diseases, using the first BBJ participants, excluding those used for PGS construction (n = 134,176; Supplementary Fig. 5). For blood chemistry tests, a linear regression analysis was performed, adjusting for age, age-squared, sex, and the first ten genetic principal components. Participants with missing values for each test were excluded from each analysis. We performed logistic regression for the onsets of T2D, CAD, and ischemic stroke. The onsets of myocardial infarction, unstable angina, and stable angina were combined as a composite endpoint to define the onset of CAD. Participants with a history of disease onset at the time of recruitment or during the follow-up period were classified as having a history of disease onset. Otherwise, they were classified as having no history of onset. In this logistic regression analysis, three sets of covariates were used. One set is the same as that used in the linear regression analysis described above (Model 1). Another set additionally includes circulating TG levels, HDL-C levels, and LDL-C levels, and BMI (Model 2). The remaining model includes SBP, DBP, smoking, and drinking in addition to the covariates in Model 2 (Model 3).

Survival analysis was performed to estimate the time to the onset of T2D, CAD, and ischemic stroke. Age was used as the time scale, with the onset as the event of interest. Participants who were censored due to no onset by the end of the follow-up period were treated as non-events in the analysis. We used the Kaplan-Meier curves to visualize the cumulative incidence across stratified three F_BCAA-Sub_ PGS groups (Top 10%: n = 13,418; Middle: n = 107,340; Bottom 10%: n = 13,418; Supplementary Table 14). The log-rank test was employed to compare survival curves among these groups. Sensitivity analyses were performed using different thresholds to define groups, 5% and 20%. All statistical analyses and visualizations were conducted using R (v.4.3.2).

## Data availability

Summary statistics for GWAS of BCAAs in BBJ1-180K are available at the GWAS Catalog (https://www.ebi.ac.uk/gwas/) under accession IDs GCSTxxx–GCSTxxx. Genotype and NMR data in BBJ1-180K were deposited at the National Bioscience Database Center (NBDC; https://humandbs.dbcls.jp/) Human Database (Genotype data in BBJ1-180K, research ID: hum0014; NMR data, research ID: hum0311). The imputation reference panel constructed using whole genome data of 3,256 Japanese individuals from BBJ and 2,504 individuals from 1000 Genome Project was also deposited at the NBDC Human Database (research ID: hum0014). The GWAS summary statistics from TMM is available at the Japanese Multi Omics Reference Panel (jMorp; https://jmorp.megabank.tohoku.ac.jp/). The GWAS summary statistics from the GWAS Catalog is available at the following URL: https://www.ebi.ac.uk/gwas/. The GWAS summary statistics from the following consortia and biobanks are publicly available at the corresponding URL: GLGC (http://www.lipidgenetics.org/), GIANT (https://portals.broadinsti-tute.org/collaboration/giant/index.php/GIANT_consortium_data_files), METSIM (https://pheweb.org/metsim-metab/), MAGIC (https://magicinvestigators.org/downloads/index.html), KoGES (https://koges.leelabsg.org/).

## Code availability

The following publicly available software and packages were used for the analyses in this study: Eagle (v.2.4.1; https://alkesgroup.broadinstitute.org/Eagle), Minimac4 (v.1.0.2; https://github.com/statgen/Minimac4), BOLT-LMM (v.2.4.1; https://alkesgroup.broadinstitute.org/BOLT-LMM), METAL (v.2011-03-25; http://csg.sph.umich.edu/abecasis/Metal/index.html), LDSC (v.1.0.1; https://github.com/bulik/ldsc), Popcorn (v.1.0.0; https://github.com/brielin/Popcorn), Genomic SEM (v.0.0.5; https://github.com/GenomicSEM/GenomicSEM), GWASLab (v.3.4.45; https://github.com/Cloufield/gwaslab), HyPrColoc (v.0.0.2; https://github.com/jrs95/hyprcoloc), PRS-CSx (v.1.0.0; https://github.com/getian107/PRScsx), R (v.4.3.2; https://www.r-project.org/).

## Data Availability

Summary statistics for the GWAS of BCAAs in BBJ1-180K will be made publicly available at the GWAS Catalog prior to or upon publication. Genotype and NMR data in BBJ1-180K were deposited at the National Bioscience Database Center (NBDC; https://humandbs.dbcls.jp/) Human Database (Genotype data in BBJ1-180K, research ID: hum0014; NMR data, research ID: hum0311). The imputation reference panel constructed using whole genome data of 3,256 Japanese individuals from BBJ and 2,504 individuals from 1000 Genome Project was also deposited at the NBDC Human Database (research ID: hum0014). The GWAS summary statistics from TMM is available at the Japanese Multi Omics Reference Panel (jMorp; https://jmorp.megabank.tohoku.ac.jp/). The GWAS summary statistics from the GWAS Catalog is available at the following URL: https://www.ebi.ac.uk/gwas/. The GWAS summary statistics from the following consortia and biobanks are publicly available at the corresponding URL: GLGC (http://www.lipidgenetics.org/), GIANT (https://portals.broadinsti-tute.org/collaboration/giant/index.php/GIANT_consortium_data_files), METSIM (https://pheweb.org/metsim-metab/), MAGIC (https://magicinvestigators.org/downloads/index.html), KoGES (https://koges.leelabsg.org/).

## Acknowledgements

We thank Peter Würtz and Machiko Usui (Nightingale Health) for their valuable support, and we acknowledge Nightingale Health Japan Plc for providing the metabolic biomarker data through a collaboration with BBJ. We also thank Chikashi Terao, Kohei Tomizuka, and Satoshi Koyama for their generous support. We are also grateful to all the participants and staff of the BBJ project. This study was supported by the Ministry of Education, Culture, Sports, Science, and Technology (MEXT) of the Japanese government and the Japan Agency for Medical Research and Development (AMED) under grant Numbers JP18km0605001 / JP23tm0624002 (the BioBank Japan project), JP19km0405215, and JP223fa627011. The super-computing resource was provided by Human Genome Center, the Institute of Medical Science, the University of Tokyo (http://sc.hgc.jp/shirokane.html).

## Author contributions

S.N., M.K. and Y.K. conceived and designed the study. S.N., Y.H. and M.K. performed the analyses. K.M., T.M., and A.N. managed the BBJ data, and T.M. led the BBJ metabolomics project. H.T. and Y.S. handled sample management. S.N. wrote the manuscript with critical input from Y.T., M.K. and Y.K. Y.K. supervised the project. All authors reviewed the manuscript and approved the final version.

