## Supplementary Figures for "Genetic influence of BCAA metabolism on type 2 diabetes and coronary artery disease, independent of traditional risk factors"

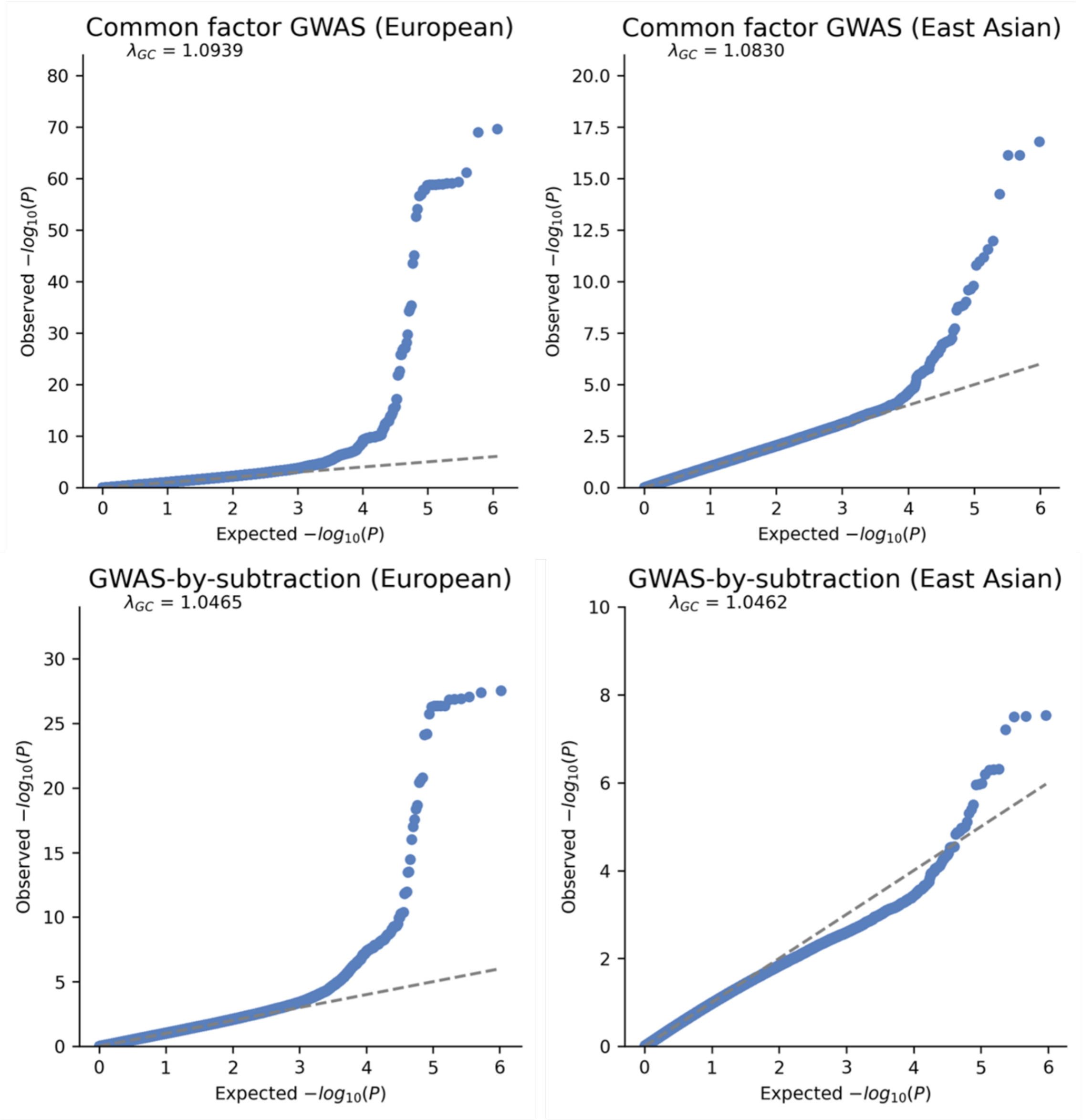


**Supplementary Fig. 1 | Quantile-quantile plots for multivariate GWASs using Genomic SEM.**

Quantile-quantile plots for common factor GWAS and GWAS-by-subtraction analyses in European and East Asian populations. The x-axis represents to expected *P* value, while the y-axis represents to the observed *P* value.

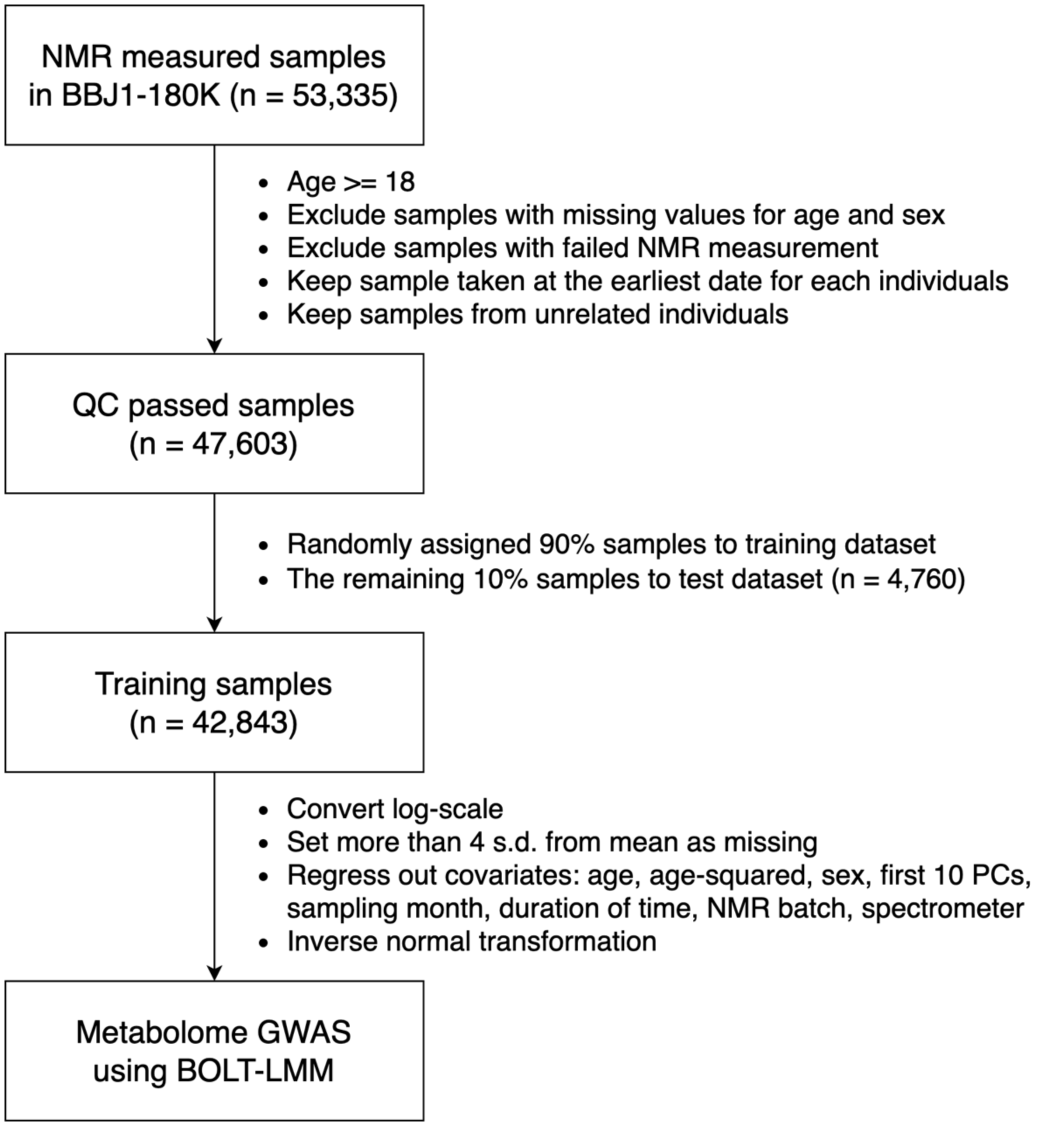


**Supplementary Fig. 2 | GWAS workflow.**

The largest GWAS for circulating BCAA levels in East Asian population was conducted using serum samples from the BBJ1-180K cohort (n = 53,335). After quality control, 47,603 samples remained. These samples were randomly divided into two datasets: a training dataset (n = 42,843) and a test dataset (n = 4,760). Samples in the training dataset were used for GWAS after transformation and adjustment.

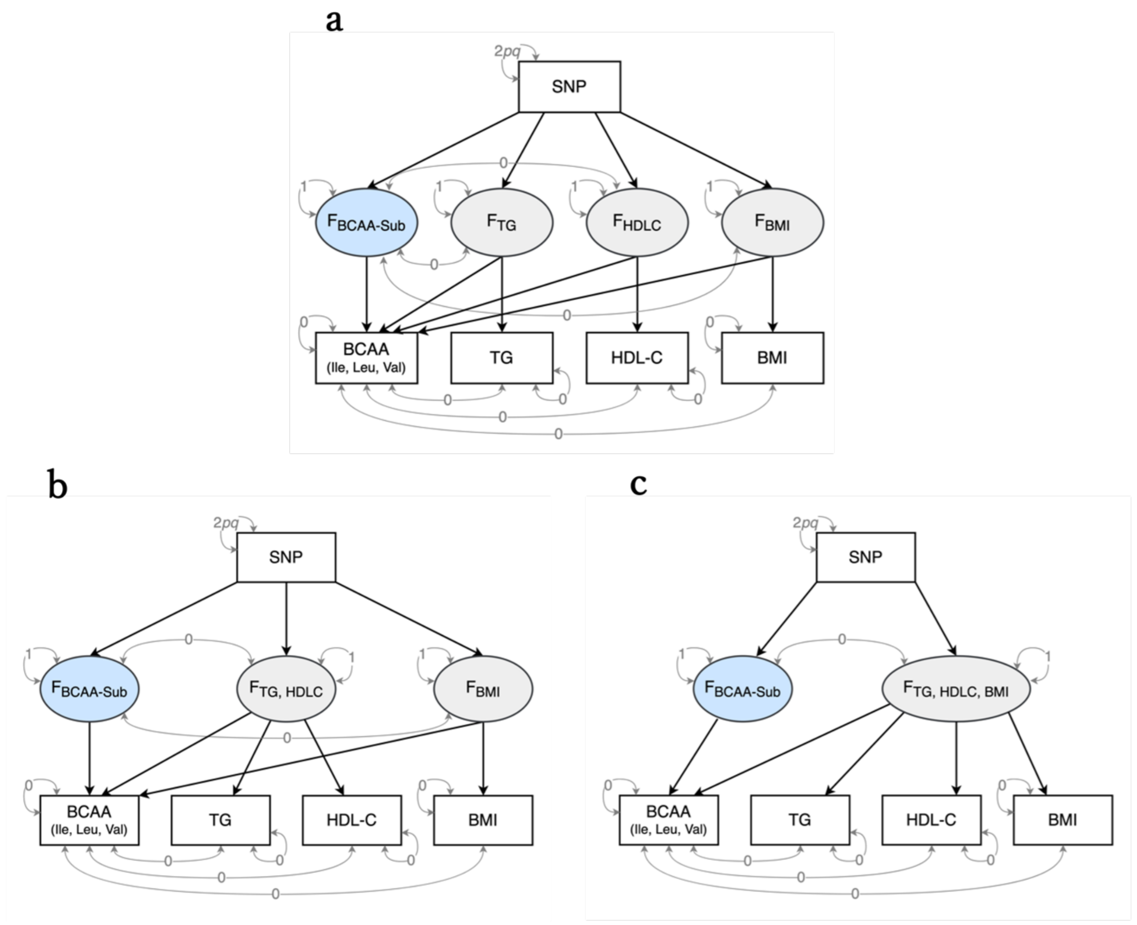


**Supplementary Fig. 3 | GWAS-by-subtraction models.**

Path diagram modeling multivariable genetic architecture. The. squares represent observed variables based on GWAS summary statistics and the circules represent latent factors. The single-headed arrows are regression relations and the double-headed arrows are variances. The variances of latent factors are fixed to 1. The variance of SNP is fixed to the value of $2pq$ ($p$ = reference allele frequency, $q$ = alternative allele frequency). The residual variances of observed variables except for SNP are fixed to 0. The covariances between the shared latent factor among BCAAs and the other latent factor(s) and between measurements of BCAAs and the other measurements are fixed to 0. **a**, The Model 1 consists of the $F_{\text{BCAA-Sub}}$ and three latent factors, each influencing not only BCAAs but also TG, HDL-C, and BMI. **b**, The Model 2 consists of three latent factors influencing BCAAs, including the $F_{\text{BCAA-Sub}}$. One of the remaining factors influences both TG and HDL-C, while the other affects BMI. **c**, The Model 3 consists of the $F_{\text{BCAA-Sub}}$ and one latent factor that influences TG, HDL-C, BMI, and BCAAs.

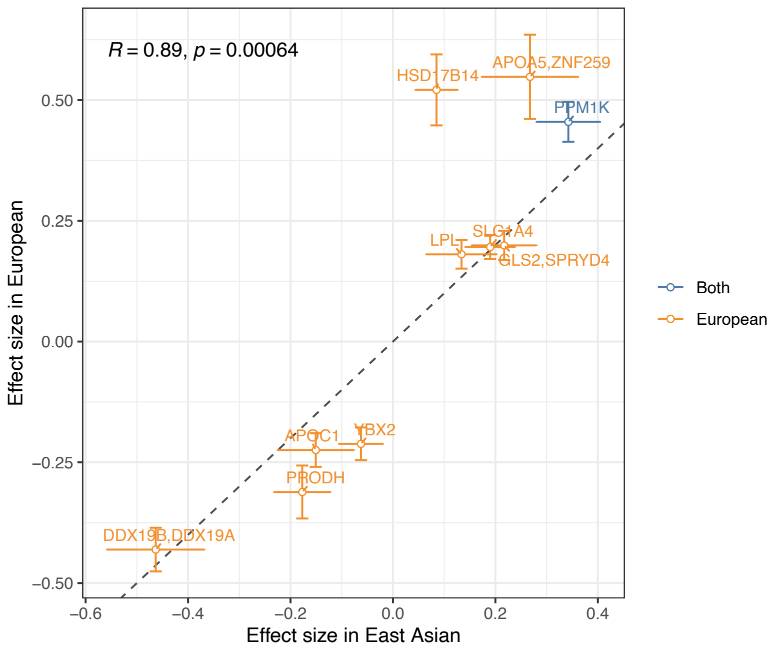


**Supplementary Fig. 4 | Comparison of cross-population effect sizes for the $F_{BCAA\text{-}Sub}$.** The scatter plot illustrates the effect sizes of lead variants for the $F_{BCAA\text{-}Sub}$. The lead variants commonly genotyped or imputed in both European and East Asian populations are visualized in this plot. The blue indicates a variant that reaches genome-wide significance ($P < 5.0 \times 10^{-8}$) in both European and East Asian populations, while the orange indicates variants that reach this level only in European population. The error bars represent standard errors. The dashed line represents y = x. Pearson's coefficient is shown in the top-right corner.

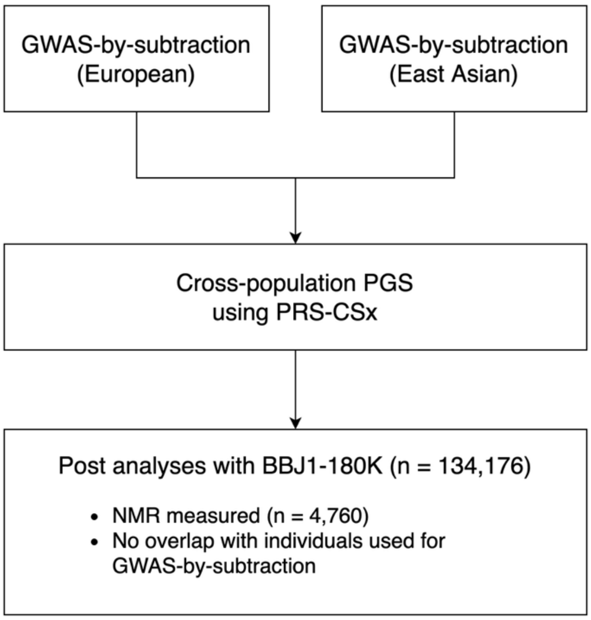


**Supplementary Fig. 5 | PGS analysis workflow.**

PGS weights for each latent factor in GWAS-by-subtraction model were constructed from their summary statistics using PRS-CSx. These PGSs were evaluated using BBJ1-180K participants who were not involved in the PGS construction. The same procedure was performed for the latent factor in the common factor model.

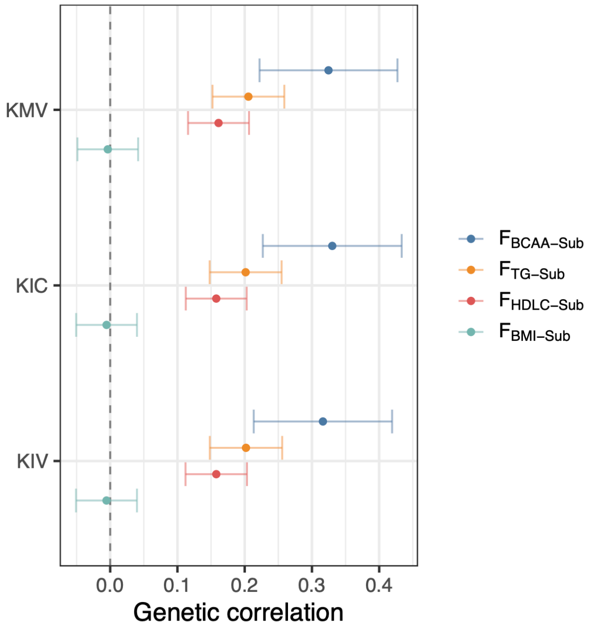


**Supplementary Fig. 6 | Genetic correlations between the latent factor in the GWAS-by-subtraction model and circulating BCKA levels.**

Estimated genetic correlations between the latent factors defined by the GWAS-by-subtraction model and circulating BCKA levels (KMV, KIC, and KIV) were visualized using summary statistics in European population. The error bars represent standard errors.

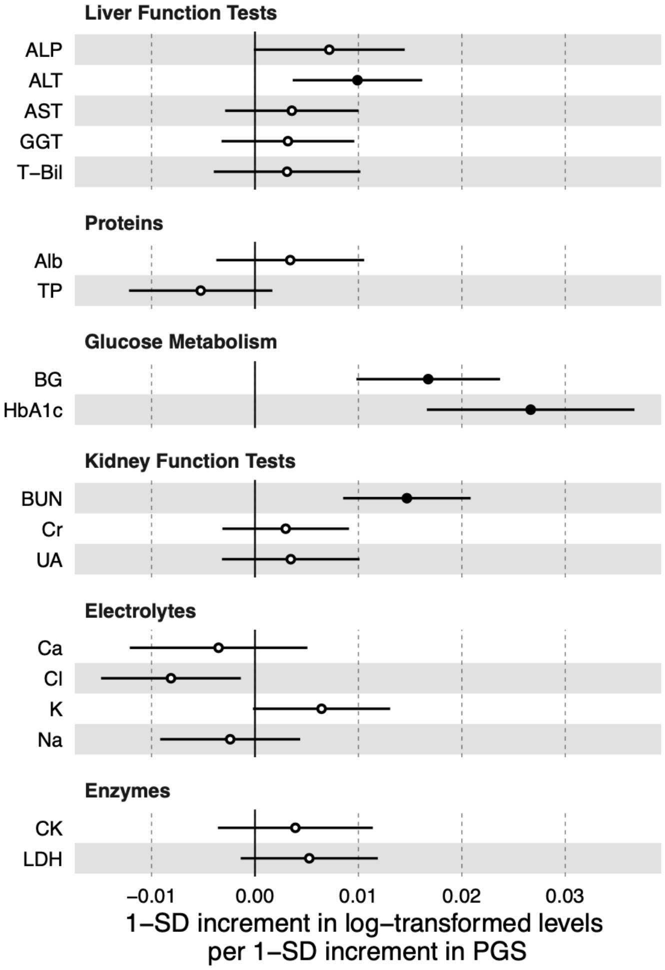


**Supplementary Fig. 7 | Associations between the $F_{BCAA\text{-}Sub}$ and 18 blood chemistry tests.** The forest plot illustrates the associations between a 1-SD increment in $F_{BCAA\text{-}Sub}$ PGS and a 1-SD increment in log-transformed levels of 18 blood chemistry tests. The error bars represent 95% CIs. The results are from linear regression analyses conducted on individuals who had never been diagnosed with T2D and were not included in PGS construction. The solid circles denote statistically significant associations (Bonferroni-corrected threshold $P <$ 0.05/18). ALP, alkaline phosphatase; ALT, alanine aminotransferase; AST, aspartate aminotransferase; GGT, gamma-glutamyl transferase; T-Bil, total bilirubin; Alb, albumin; TP, total protein; BG, blood glucose; HbA1c, hemoglobin A1c; BUN, blood urea nitrogen; Cr, creatinine; UA, uric acid; Ca, calcium; Cl, chloride; K, potassium; Na, sodium; CK, creatine kinase; LDH, lactate dehydrogenase.

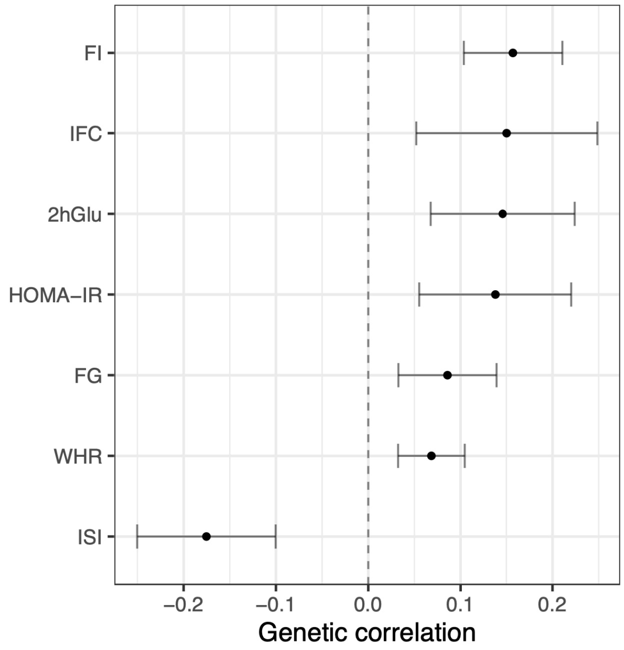


**Supplementary Fig. 8 | Genetic correlations between the $F_{BCAA\text{-}Sub}$ and IR-related traits.** Genetic correlations between $F_{BCAA\text{-}Sub}$ and six IR-related traits adjusted for BMI. The points represent estimated genetic correlations. The error bars indicate standard errors.

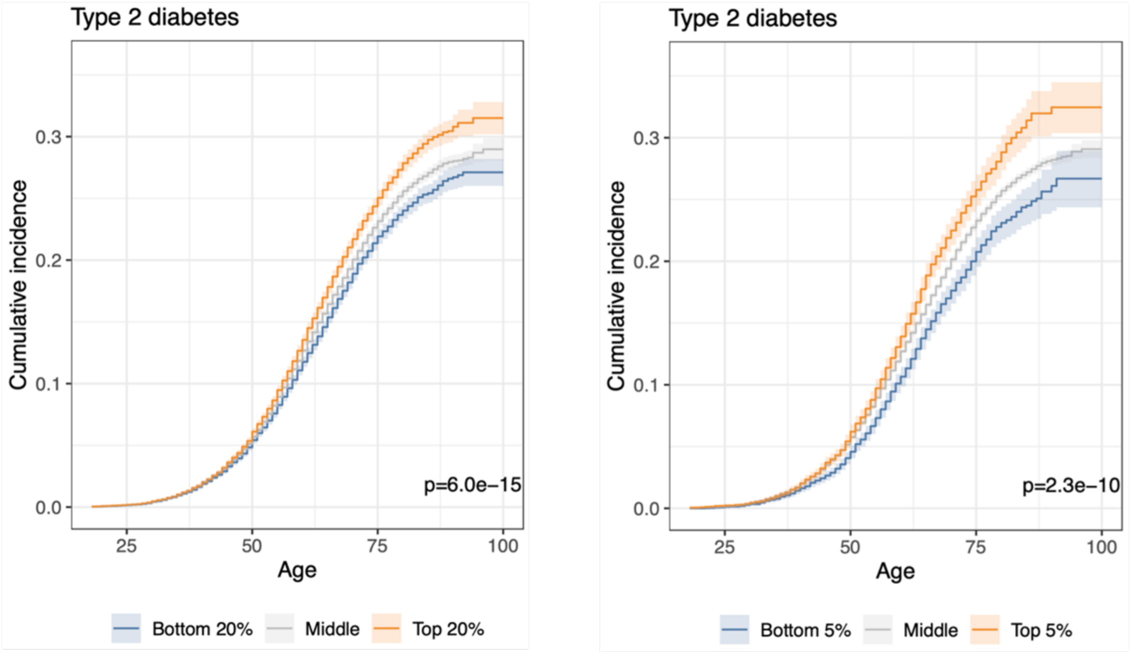


**Supplementary Fig. 9 | Cumulative incidence curves for the onset of T2D by $F_{BCAA\text{-}Sub}$ PGS strata.**

The Kaplan-Meier curves display the cumulative incidence across three stratified $F_{BCAA\text{-}Sub}$ PGS groups defined by different thresholds. The left panel shows the top and bottom 20%, while the right panel shows the top and bottom 5%. The *P* values from the log-rank tests are displayed in the bottom-right corner.

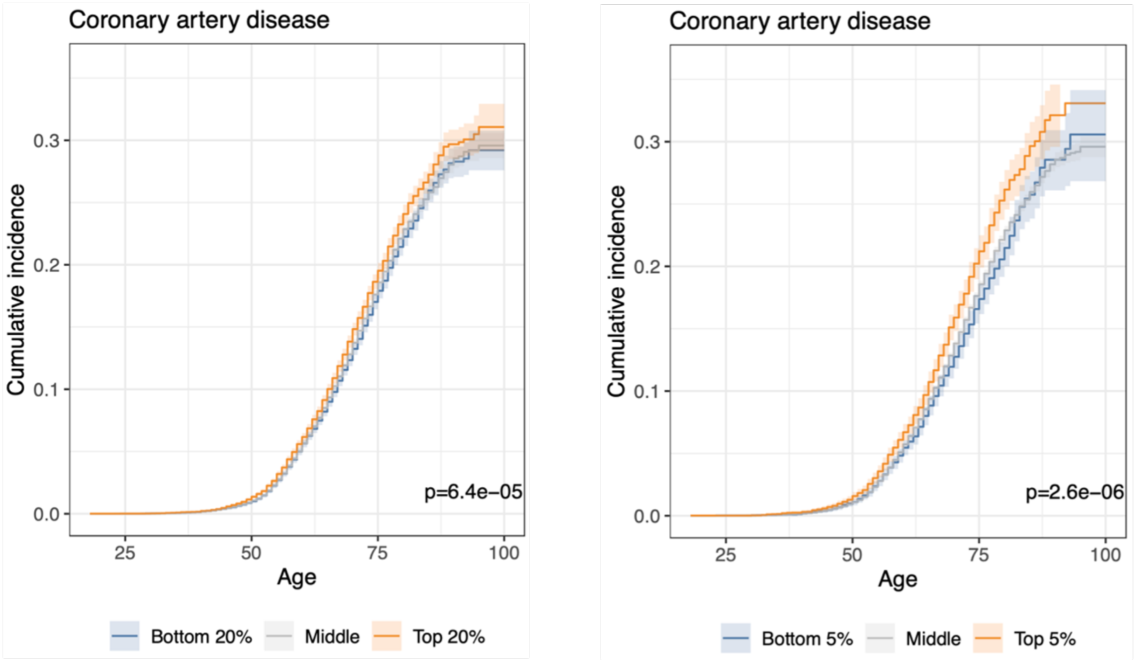


**Supplementary Fig. 10 | Cumulative incidence curves for the onset of CAD by $F_{BCAA\text{-}Sub}$ PGS strata.**

The Kaplan-Meier curves display the cumulative incidence across three stratified $F_{BCAA\text{-}Sub}$ PGS groups defined by different thresholds. The left panel shows the top and bottom 20%, while the right panel shows the top and bottom 5%. The *P* values from the log-rank tests are displayed in the bottom-right corner.

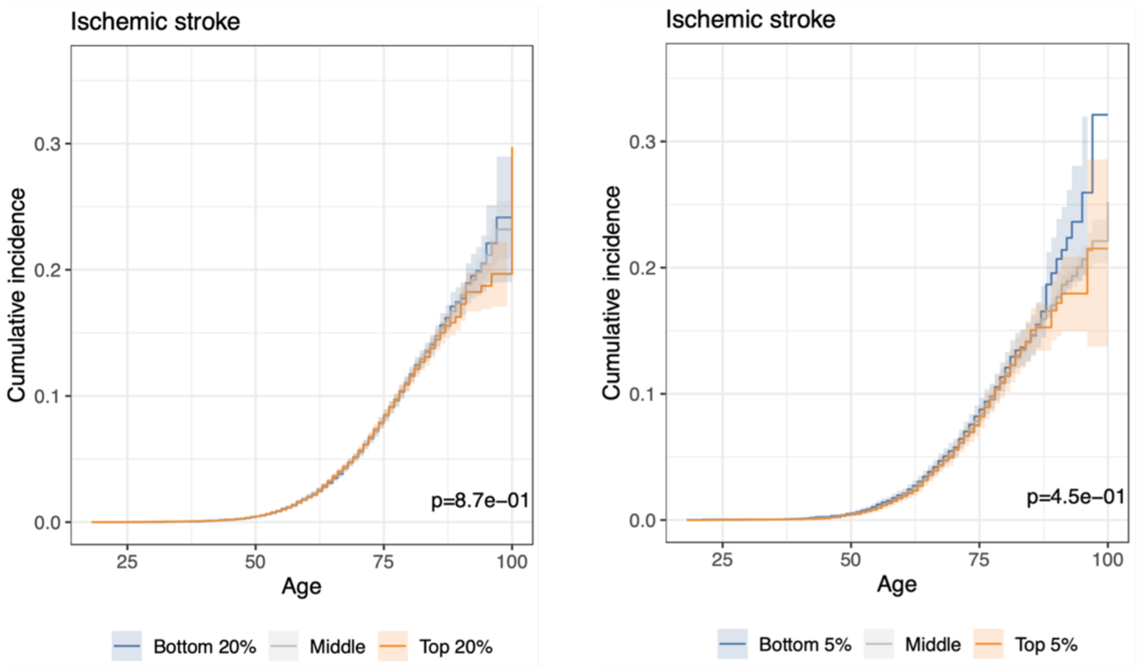


**Supplementary Fig. 11 | Cumulative incidence curves for the onset of ischemic stroke by $F_{BCAA\text{-}Sub}$ PGS strata.**

The Kaplan-Meier curves display the cumulative incidence across three stratified $F_{BCAA\text{-}Sub}$ PGS groups defined by different thresholds. The left panel shows the top and bottom 20%, while the right panel shows the top and bottom 5%. The *P* values from the log-rank tests are displayed in the bottom-right corner.
